# Physician-assisted suicide among older adults in Austria (2022-2023): Sociodemographic and comorbidity profile

**DOI:** 10.64898/2026.07.09.26357607

**Authors:** Emilise Lucerne Pötz, Anna Schultz, Christian Jagsch, Carlos Watzka, Wolfgang Freidl, Erwin Stolz

## Abstract

In 2022, Austria legalised physician-assisted suicide (PAS). Among 1,847,919 older adults, there were 92 PAS and 977 unassisted suicides (UAS) in 2022-2023. Compared to the general population, older adults who died by either PAS or UAS, were older and more likely to live alone. Compared with UAS, older adults who died by PAS were more likely to be female, higher-educated, live in urban areas, and diagnosed with cancer, or diseases of the nervous system, and less likely diagnosed with mental/ behavioural disorders. PAS and UAS among older adults in Austria showed different sociodemographic and comorbidity characteristics.

## INTRODUCTION

In January 2022, Austria enacted the Assisted Dying Act (“Sterbeverfügungsgesetz”)[^1^], legalising physician-assisted suicide (PAS) for adults with a terminal illness or severe permanent condition causing persistent impairments in daily life. Potential applicants must be of legal age and possess decision-making capacity and personally draft a formal directive (“Sterbeverfügung”) stating their wish to end their life. Two physicians, one of whom must be a palliative care physician, are required to inform the applicant about the procedure and confirm the presence of the required conditions. Then, after a waiting period for 12 weeks (2 weeks in terminal cases), the lethal substance can be obtained.[^2^] Euthanasia, that is, non-self-administered medical aid in dying (MAID) remains illegal in Austria.

Older adults aged 65 years and above constitute a significant proportion of those who die by PAS[^3^] and at the same time have the highest rates of unassisted suicide (UAS) out of all age groups[^4–7^]. Both PAS and UAS are associated with poor health and are socio-demographically patterned, but direct comparisons remain sparse. A single study[^3^] from Switzerland found UAS to be associated with male gender and lower education, while PAS was associated with higher education. Both PAS and UAS were associated with no religious affiliation, living alone, and having no children. With regard to comorbidities, separate studies found major depression, cancer, liver disease, cerebrovascular disease, CODP, arthritis, prostate and male genital disorders, and spinal fractures to be associated with UAS[^8–11^], whereas cancer, diseases of the nervous, circulatory, and respiratory system have been associated with PAS[^3,12^].

The aim of this research is to bridge this gap in the research literature and to add to our understanding of PAS and UAS and their risk factors in older adults, leveraging newly available individual-level register data from Austria.

## METHODS

### Data

In this retrospective cohort study, we used data from several national population registers (Central Population Register, Central Civil Status Register, and Register-based Labour Market Statistics) via the Austrian Micro Data Center (AMDC). The study population consisted of all Austrians aged 65+ who were alive at the beginning of 2022.

### Variables

Suicide deaths were determined based on the 10^th^ edition of the International Classification of Disease (ICD-10) codes X60-X84, Y87.0. PAS was identified from the Austria-specific code U060 (“assisted suicide”). Sociodemographic variables included gender (male/female), age (65-74/75-84/85+ years), education (primary/secondary/tertiary), living alone (no/yes), area of living (urban/intermediate/rural), and place of birth (Austria/other country). Comorbidities at the time of death were categorized as cancer (ICD-10:C00-C97), mental/behavioural disorders (ICD-10:F00-F99), diseases of the endocrine/nutritional/metabolic (ICD-10:E00-E90), nervous (ICD-10:G00-G99), circulatory (ICD-10:I00-I99), and respiratory (ICD-10:J00-J99) system.

### Statistical analysis

We calculated cumulative incidences of PAS and UAS per 100,000 persons based on follow-up from 1^st^ of January 2022 until the date of death or end of study (31^st^ of December 2023). Odds ratios (OR) for sociodemographic factors and comorbidities comparing PAS and UAS were estimated from a logistic regression model[^13^] appropriate for rare events using R-package *logistf* (v1.26.1). All calculations were conducted with R (v4.4.3).

## RESULTS

In 2022-2023, there were 92 cases of PAS and 977 cases of UAS among 1,847,919 older Austrians and a cumulative incidence of 5 respectively 53 for PAS and UAS per 100,000 persons (Table 1). Compared with the total older population, individuals who died by either PAS or UAS were older and more likely to live alone. Those dying by PAS had more education and were more likely to live in urban areas, whereas those who died by UAS were more likely to be male, less educated, and living in rural areas.

**Table 1:** Sociodemographic characteristics of the general older population, PAS and UAS (2022-2023).

| Characteristic |  | Study population | PAS |  | UAS |  |
| --- | --- | --- | --- | --- | --- | --- |
|  |  | *N* (%) | *n* (%) | Cumulative incidence (per 100,000) | *n* (%) | Cumulative incidence (per 100,000) |
| Total (%) |  | 1,847,919 | 92 | 4.98 | 977 | 52.9 |
| Gender | Male | 810,667 (43.9) | 33 (35.9) | 4.07 | 754 (77.2) | 93 |
|  | Female | 1,037,252 (56.1) | 59 (64.1) | 5.69 | 223 (22.8) | 21.5 |
| Age | 68-74 | 1,009,003 (54.6) | 33 (35.9) | 3.27 | 407 (41.07) | 40.3 |
|  | 75-84 | 620,906 (33.6) | 44 (47.8) | 7.09 | 397 (40.6) | 63.9 |
| Education | 85+ | 218,010 (11.8) | 15 (16.3) | 6.88 | 173 (17.7) | 79.4 |
|  | Primary | 577,961 (31.3) | 12 (13.0) | 2.08 | 281 (28.8) | 48.6 |
|  | Secondary | 1,087,832 (58.9) | 59 (64.1) | 5.42 | 624 (63.9) | 57.4 |
| Living alone | Tertiary | 182,126 (9.9) | 21 (22.8) | 11.5 | 72 (7.4) | 39.5 |
|  | Yes | 588,458 (31.8) | 40 (43.5) | 6.8 | 394 (40.3) | 69 |
|  | No | 1,259,461 (68.2) | 52 (56.5) | 4.13 | 583 (59.7) | 66.3 |
| Area of living | Urban | 561,361 (30.4) | 49 (53.3) | 8.73 | 231 (23.6) | 41.1 |
|  | Intermediate | 586,833 (31.8) | 24 (26.1) | 4.09 | 319 (32.7) | 54.4 |
|  | Rural | 699,725 (31.8) | 19 (20.7) | 2.72 | 427 (43.7) | 61 |
| Place of birth | Austria | 1,571,880 (85.1) | 76 (82.6) | 4.83 | 904 (92.5) | 57.5 |
|  | Other | 276,039 (14.9) | 16 (19.51) | 5.8 | 73 (7.5) | 26.4 |

Note. *N* = 1,847,919, PAS = physician-assisted suicide, UAS=unassisted suicide.

Results from the adjusted logistic regression model (Supplementary Table 1) again showed that compared to UAS, individuals who died by PAS were more likely female (OR=9.31, 95%-CI=5.09,17.72), higher educated (tertiary vs. primary education: OR=4.92, 95%-CI=1.90,13.15), and lived in urban areas (rural vs. urban: OR=0.35, 0.17,0.71). The most common comorbidities in PAS were cancer (*n*=48; 52%), diseases of the circulatory (*n*=31;34%), nervous (*n*=25;27%), and respiratory system (*n*=13;14%). Among UAS the most common comorbidities were mental/behavioural disorders (*n*=225;23%), diseases of the circulatory system (*n*=159;20%), and cancer (*n*=104;11%). In comparison to UAS, older adults who died by PAS were far more likely to have been diagnosed with cancer (OR=18.45, 95%-CI=9.96,35.61) and diseases of the nervous system (OR = 12.58, 95%-CI=5.86,27.58), and far less likely to be diagnosed with a mental/behavioural disorders (OR=0.24, 95%-CI=0.09,0.54).

## DISCUSSION

Austria legalised PAS in 2022.[^1^] Among countries that have legalised MAID, which often includes PAS, legal requirements vary widely – particularly regarding eligibility of minors, individuals with psychiatric conditions, or those without terminal illness – with Austria taking a restrictive stance.[^14,15^] In this study, we compared characteristics of older adults who died by PAS in Austria with those who died by UAS; two forms of suicide that have largely been treated as separate issues in both the scientific literature and public discourse. Compared to UAS, those who died by PAS, were more often female, higher educated, urban-dwelling, and diagnosed with cancer or nervous system diseases, and less likely to have mental/behavioural disorders.

Suicides in old age tend to be less impulsive, marked by higher suicidal intent and are more lethal than in younger age groups[^16,17^], which suggests ongoing suffering and the wish to no longer continue living under burdensome conditions. While PAS requires navigating a complex formal approval process unlike UAS, both forms of suicide are closely linked to severe illness and reduced quality of life in older adults.[^3,8^]. A cancer diagnosis, for example, is a known risk factor for suicide[^18^], and featured as a comorbidity in our study in both UAS and PAS, but much more heavily so in the latter, which is in line with previous research[^3,19,20^]. This can be linked to the well-prognosticated limited life expectancy[^21^] and low quality of life[^22^] of many cancer patients. In contrast, diagnoses of a mental/behavioural disorders were prevalent only in UAS, which again is line with previous research[^23–26^]; UAS pre- and postvention should put particular emphasis on reaching older men with low education levels residing in rural areas. Among PAS, mental/behavioural disorders were rare, which can be attributed to Austrian law[1] requiring both a terminal or incurable disease and consistent decision-making capacity, which renders PAS solely due to mental/behavioural disorders such as depression highly unlikely at the current time.

In keeping with the robust body of evidence on UAS for both younger and older age groups, we also observed higher UAS rates among men compared to women.[^9,27–29^] For PAS, research from Switzerland[^3,19^] also reported higher rates in women, similarly to research from the Netherlands, Belgium, and Canada[^30–32^]. Another study[^20^] from Switzerland found higher rates of PAS in women until age 69, with men having have higher rates thereafter, which may have to do with the long tradition of legal PAS in that country. Meta-analytic evidence shows women tend to seek out their general practitioner more frequently and have more positive attitudes towards seeking psychological help.[^34,35^] The willingness to seek out care might explain, in part, the tendency to seek out PAS as opposed to UAS. Moreover, there is extensive evidence that men show higher average aggression and more often display aggressive behaviour than women.[^36,37^] These differences might account for women to more frequently use PAS than UAS – the latter being an often violent and solitary act. Similar to a previous study[^3^], we also found higher education associated with PAS, while lower education was associated with UAS. Individuals with higher education might be better informed about PAS and put more trust in the bureaucratic and health care system compared to those with more limited education. Our results suggest that living with others may protect against both forms of suicide, which also aligns with previous work[^38–42^]. Urban residence was associated with PAS and rural residence with UAS, which is consistent with previous research[^43,44^] in Austria and elsewhere[^45^], and might be due to better access to healthcare (e.g. higher number of doctors or specialized doctors per person) and more liberal attitudes towards MAID in urban areas[^46^]. These differential patterns could reflect differences in knowledge regarding available end-of-life care, as well as sociocultural differences pertaining to liberalism, individualism, and gender norms. In other words, when facing severe illness, older individuals may have similar motivations to end their lives, but they may pursue different paths to do so depending on socio-cultural and contextual factors.

This is the first empirical study of PAS in Austria based on individual-level register data and one of only two studies explicitly comparing characteristics of PAS and UAS cases. A limitation of this study is the small number of PAS among older adults in Austria so far, which limited the number of sociodemographic and comorbidity characteristics that could be explored.

## Conclusion

During the first two years after legalisation, PAS in older adults was predominated by women, those with higher education, and those living in urban areas. In contrast, the ten-fold higher number of UAS are dominated by older men in rural areas with low education. Future research should monitor trends in PAS in Austria closely and attempt to shed light on the socio-cultural mechanisms shaping the differential between UAS and PAS among older adults.

---

**Supplementary Table 1:** Sociodemographic variables and comorbidities in PAS compared to UAS (2022-2023).

| Variable | OR (95% CI) |
| --- | --- |
| Female gender (ref.: male) | 9.31 (5.09-17.72) |
| 75-84 years (ref.: 65-74 years) | 1.20 (0.66-2.19) |
| 85+ years (ref.: 65-74 years) | 1.62 (0.71-3.61) |
| Secondary education (ref.: primary education) | 2.50 (1.22-5.50) |
| Tertiary education (ref.: primary education) | 4.92 (1.90-13.15) |
| ≥ 2 person household (ref.: living alone) | 1.17 (0.66-2.07) |
| Intermediate (ref.: urban area of living) | 0.62 (0.31-1.20) |
| Rural (ref.: urban area of living) | 0.35 (0.17-0.71) |
| Born in Austria (ref.: born in other country) | 0.74 (0.34-1.69) |
| Cancer (ref.: no) | 18.45 (9.96-35.61) |
| Endocrine, nutritional and metabolic disease (ref.: no) | 2.45 (0.85-6.63) |
| Mental and behavioural disorder (ref.: no) | 0.24 (0.09-0.54) |
| Nervous system disease (G00-G99) (ref.: no) | 12.58 (5.86-27.58) |
| Circulatory system disease (I00-I99) (ref.: no) | 1.55 (0.77-3.10) |
| Respiratory system disease (J00-J99) (ref.: no) | 2.37 (0.99-5.43) |

*Note.* $N=1,069$ ; 92 cases of physician-assisted suicide, 977 cases of unassisted suicide, results based on logistic regression analysis with Firth correction; PAS = physician-assisted suicide, UAS=unassisted suicide, 95%-CI = 95% confidence interval.

## Patient consent for publication

Not applicable.

## Ethics approval

The conduct of this study was approved by the Ethics Committee of the Medical University of Graz (1172/2024).

## Data

Access and linkage of register data (Central Population Register, Central Civil Status Register, Cancer Register, and Register-based Labour Market Statistics) was approved of and provided by the Austrian Micro Data Center (AMDC) of Statistics Austria following national legislation (Bundesstatistikgesetz §31, Forschungsorganisationsgesetz §38b). The AMDC is a research data infrastructure facility of Statistics Austria that enables research on micro data processed in compliance with data protection regulations. The data used for this research can be accessed by researchers at scientific institutions accredited with the AMDC against a fee. For further information, see https://www.statistik.at/en/services/tools/services/center-for-science/austrian-micro-data-center-amdc.

